# Age and body-mass index account for part of the sex differences in cerebral venous thrombosis

**DOI:** 10.64898/2026.11.11.26367881

**Authors:** Tiziana Ielapi-Giuffrida, Marah Ruepp, Lisa Herzog, Meret Branscheidt, Mira Katan, Patrick Thurner, Zsolt Kulcsar, Andreas Luft, Michael Weller, Susanne Wegener, Anton Schmick

**Affiliations:** Department of Neurology, University Hospital and University of Zurich, Zurich, Switzerland; D-HEST, ETH Zurich, Switzerland; Cereneo Center for Neurology and Rehabilitation, Vitznau, Switzerland; Department of Neurology, University of Basel, Basel, Switzerland; Department of Neuroradiology, University Hospital of Zurich, Zurich, Switzerland

**Author notes:** Corresponding author: Dr. med. Anton Schmick, Department of Neurology, University Hospital Zurich, Frauenklinikstrasse 26, 8091 Zurich, Switzerland.

**Keywords:** cerebral venous thrombosis, sex differences, headache, inflammation, platelets, functional outcome

## Abstract

**Background:** Women with cerebral venous thrombosis are younger than men and more often present with headache, but these differences have not been adjusted for age or body-mass index, both of which differ by sex.

**Methods:** Single-center cohort of 219 consecutive adults with confirmed cerebral venous thrombosis admitted in Zurich in 2001-2021. Presentation, imaging and laboratory findings were compared between women and men, and every difference was re-estimated at equal age and body-mass index. Secondary analyses compared functional outcome, residual symptoms and return to work by sex, with an audit of incomplete follow-up.

**Results:** Women (132, 60.3%) were younger (47.3 versus 54.9 years) and had a lower body-mass index (25.9 versus 27.8 kg/m²) than men. Headache (72.0% versus 57.5%) and cortical vein thrombosis (14.4% versus 5.7%) were more frequent in women, but neither association persisted after adjustment (odds ratios, 1.17 [95% CI, 0.54-2.54] and 2.05 [95% CI, 0.59-7.06]). Higher platelet counts (+56 × 10⁹/L [95% CI, 26-86]) and elevated acute-phase markers without documented infection (59.1% versus 33.3%; odds ratio, 2.91 [95% CI, 1.44-5.87]) persisted after adjustment for age, body-mass index and calendar period. Among assessed patients (38% at 6 months), functional independence (86.3% versus 87.5%), residual headache, return to work and recanalization did not differ by sex.

**Conclusions:** At equal age and body-mass index, the female excess of headache and cortical vein involvement was no longer apparent, whereas a higher platelet count and a non-specific acute-phase response remained. Neither translated into a difference in outcome among assessed patients.

**Clinical Perspective:** *What Is New?:*

- In 219 consecutive adults with cerebral venous thrombosis, the female excess of headache and cortical vein thrombosis reported by the large registries was attenuated once age and body-mass index were accounted for, whereas higher platelet counts and more frequent elevation of acute-phase markers without infection in women persisted.
- Women without a sex-specific risk factor resembled men in age and in the frequency of headache, which suggests that part of the female phenotype of cerebral venous thrombosis is a phenotype of young age and reproductive exposures.

*What Are the Clinical Implications?:*

- Sex differences in presentation were not accompanied by differences in functional outcome, residual symptoms, return to work, or recanalization among the patients assessed, so sex alone does not identify patients who need a different follow-up.

## Introduction

Cerebral venous thrombosis (CVT) is more common in women than in men, and the sex ratio has been attributed largely to oral contraception, pregnancy and the puerperium.^1–3^ Whether being a woman also changes the course of the disease is less clear. In the International Study on Cerebral Vein and Dural Sinus Thrombosis (ISCVT), women had a better functional outcome than men, but the advantage was confined to women with a sex-specific risk factor.^4^ The CEVETIS cohort found no sex difference in functional outcome or mortality, and recurrent venous thromboembolism at other sites was more frequent in men.^5^ A Swedish long-term study associated female sex with not returning to work.^6^ Sex differences in outcome after CVT therefore remain insufficiently characterized.

Sex differences in presentation are better documented but harder to interpret. In ISCVT, VENOST and CEVETIS, women were younger than men, more often presented with headache (ISCVT) or seizures (VENOST), and less often had infection-associated CVT.^4,5,7^ Women with CVT are about nine years younger than men,^8^ older patients less often have headache and more often have a poor outcome,^9^ and obesity is a risk factor for CVT in women using oral contraceptives.^10^ Age and body-mass index (BMI) are thus plausible explanations for part of the reported differences, and the ISCVT investigators themselves considered the higher frequency of headache in women with a sex-specific risk factor a possible effect of age.^4^ None of these cohorts adjusted the sex comparison for age or BMI. The most recent large registry, from Vellore, documented a shift toward male predominance as postpartum CVT declined and the mean age at presentation rose, which illustrates how closely the sex ratio of a CVT cohort depends on reproductive exposures and era.^11^

Mechanistic explanations have been proposed for a female phenotype of CVT, among them a hormonal prothrombotic state, higher platelet counts and a stronger acute-phase response,^12,13^ but these have not been examined with adjustment for the demographic differences between the sexes.

Our aims were to test whether sex differences in the presentation of CVT persist after accounting for age and BMI, and to compare functional outcome, residual symptoms and occupational recovery between women and men in a cohort treated in routine care. Because follow-up in a retrospective registry is incomplete, we also compared assessed with unassessed patients and quantified how far missing outcomes could alter the conclusions.

## Methods

### Study design and population

This retrospective cohort study included consecutive patients with CVT treated at University Hospital Zurich, a tertiary academic center, and is reported according to STROBE.^14^ Cases were identified by automated searches of the electronic health records of the Department of Neurology for the ICD codes and search terms listed in the Supplementary methods, and confirmed by CT or MR venography. Inclusion criteria were age ≥18 years, admission between 1 January 2001 and 31 December 2021, and confirmed thrombosis of a dural sinus, a deep cerebral vein or a cortical vein. Traumatic CVT was not included. The study adhered to the Declaration of Helsinki and was approved by the Ethics Committee of the Canton of Zurich (KEK 2021-01558); all patients had provided general consent. Sex was abstracted from the medical records as sex assigned at birth; gender identity was not recorded (SAGER guidelines).^15^

### Data collection and definitions

Demographics, substance use, previous venous thromboembolism, hormonal exposure, thrombophilia, malignancy, D-dimer and platelet counts were abstracted from the records. Oral contraception denotes any oral contraceptive at the time of the CVT; a second field records the preparation where it was documented. Hormonal therapy denotes any other hormonal treatment, including corticosteroids, hormone replacement and levothyroxine, and is listed separately from contraception. A sex-specific risk factor in women was defined as oral contraception, pregnancy, the puerperium or estrogen or progestogen therapy; corticosteroids and levothyroxine, which the registry records in the same field, were not counted. Cancer denotes any tumor recorded as active at admission. Systemic inflammation was a composite that was positive if any of the following applied at presentation: C-reactive protein (CRP) >5 mg/L, leukocyte count above the laboratory reference range, fever ≥38.0 °C, or a clinically documented infection of any site; it was not restricted to infection-associated CVT. A free-text source field separated patients with elevated markers and no infectious focus (markers-only phenotype) from patients with a documented infection. The registry records markers as elevated or not elevated. The same rule applied to presenting symptoms and signs, which the registry codes as present or absent without a blank. The number of patients with each item documented, explicitly coded as not known, or left blank is given by sex in Supplementary Table 1.

Imaging variables were the thrombosed sinuses and veins, counted per patient as the number of sites (superior sagittal, transverse and sigmoid sinus, internal and external jugular vein, cortical veins, deep venous system); no patient had external jugular vein thrombosis. Venous infarction denotes a parenchymal lesion of non-arterial distribution attributed to the venous occlusion in the imaging reports; congestive intracerebral hemorrhage denotes any parenchymal hemorrhage attributed to the CVT, including hemorrhagic transformation of a venous infarct. Hemorrhage volume and the distinction between cytotoxic and vasogenic edema were not recorded. Clinical variables were headache and its accompanying symptoms, focal deficits, acute symptomatic seizures (≤7 days),^16^ Glasgow Coma Scale (GCS), modified Rankin Scale (mRS) and National Institutes of Health Stroke Scale (NIHSS) at admission, anticoagulation and antiseizure medication (ASM).

### Outcomes

At scheduled follow-up visits (typically 3, 6 and 12 months) the mRS, NIHSS, headache, seizures, recurrence and recanalization were recorded. Return to work was documented in patients employed before the CVT. The registry holds no date for this item, nor for persistent headache or ASM at follow-up; these items refer to the first follow-up at which they were documented. Comorbidities were abstracted from the admission letters into a fixed list. Deaths were ascertained from the hospital record; two deaths that became known from another institution are annotated in the registry (Supplementary Table 2). Recanalization was abstracted from the neuroradiology reports of the scheduled follow-up imaging as complete, partial or absent. The registry does not separate an absent from an unknown result and the reports were not re-read for this study, so recanalization by sex is a secondary, descriptive comparison (Supplementary methods).

### Statistical analysis

Categorical variables are summarized as frequencies and percentages with their denominators, and continuous variables as mean (SD) or median [IQR or range]. Women and men were compared with the chi-squared or Fisher exact test and the t-test or Mann-Whitney U test; P<0.05 was considered significant. The sex comparisons of presentation were the primary analysis and are reported without adjustment for multiple testing; Benjamini-Hochberg-adjusted P values are given in Supplementary Table 3.

Every sex difference in a clinical, imaging or laboratory feature or in an outcome was reestimated in logistic or linear regression models adjusted for age, and for age and BMI. Outcome models were additionally adjusted for admission mRS. The age- and BMI-adjusted models were fitted twice: in patients with BMI recorded, and in all patients with BMI imputed by chained equations (30 datasets, predictive mean matching from age, sex, admission severity, presentation and follow-up variables; Rubin’s rules); the same imputation supplied the admission mRS of the two patients in whom it was missing, so the imputed outcome models include one patient more than the age-adjusted models. The models were repeated with calendar period (admission before or from 2012) added, and the headache, imaging and outcome models with the number of thrombosed sites added.

Follow-up was incomplete. For each follow-up outcome, assessed and unassessed patients were compared and the probability of assessment was modeled by logistic regression. The whole-cohort proportion with mRS 0-1 at 6 months was estimated by multiple imputation (patients known to have died before the visit assigned mRS 6), by inverse-probability weighting and as extreme-case bounds. A delta-adjusted analysis subtracted 10 to 40 percentage points from the imputed probability of independence of each unassessed survivor. Time to recanalization by sex was estimated with Kaplan-Meier methods among imaged patients, with the event dated to the scheduled visit at which it was first documented and observation of the remaining patients ended at the last documented imaging assessment. Women and men were compared by the log-rank test, and the hazard ratio (HR) for women was estimated by Cox proportional-hazards regression adjusted for age and BMI, with BMI recorded and with BMI imputed. Analyses were performed with R 4.5.3.

## Results

### Cohort and documentation

From January 2001 to December 2021 the automated searches identified 2198 entries with possible non-traumatic CVT; after exclusion of miscoded and duplicate entries, 233 patients had a diagnosis of CVT. After exclusion of 8 patients without confirmatory venography, 5 aged under 18 years and one admitted before the study period, 219 patients were analyzed (Supplementary Figure 1), 132 (60.3%) of them women. Mean age was 50.3 ± 18.8 years (range 18-92).

Headache (145/219, 66.2%) and focal neurological deficits (140/219, 63.9%) were the most frequent presentations; 73 patients (33.3%) had acute symptomatic seizures. Venous infarction was present in 69 (31.5%) and congestive intracerebral hemorrhage in 77 (35.2%). Whole-cohort descriptive data are given in Supplementary Tables 4 and 5.

Presenting symptoms, parenchymal lesions, previous thromboembolism, cancer and the inflammation composite were coded for every patient, with no blank entries. Laboratory and anthropometric items were incomplete: a D-dimer result was documented in 62.1% of women and 69.0% of men, BMI in 65.9% and 75.9% (P=0.12), smoking status in 76.5% and 75.9%, and a platelet count in 98.5% and 100% (Supplementary Table 1). Documentation did not differ between the sexes for any item.

### Demographics, exposures and risk factors

Women were younger than men (47.3 ± 19.6 versus 54.9 ± 16.5 years; P=0.002) and had a lower BMI (25.9 ± 6.1 versus 27.8 ± 6.4 kg/m²; P=0.034; Table 1). Of the 132 women, 52 (39.4%) used oral contraception (an estrogen-containing preparation in 24, a progestogen-only preparation in 4, preparation not specified in 24), 2 (1.5%) were pregnant, 3 (2.3%) were postpartum and 16 (12.1%) received other hormonal therapy. In all, 63 women (47.7%) had at least one sex-specific risk factor (an estrogen or progestogen preparation in 6 of the 16 on hormonal therapy) and 69 (52.3%) had none. Four men (4.6%) received hormonal therapy. Women with a sex-specific risk factor were younger (mean 34.1 years) than women without (59.4 years) and than men (54.9 years) (Supplementary Table 6). Men more often had a history of venous thromboembolism (24.1% versus 13.6%; P=0.047) and more often received lifelong anticoagulation (13.8% versus 2.3%; P=0.002). Cancer, known hematologic disease and chronic inflammatory or autoimmune disease did not differ by sex (Table 1). More women than men had been admitted before 2012 (41.7% versus 18.4%).

**Table 1.** Presentation, imaging, laboratory findings and treatment by sex (unadjusted)

|  | Women (n = 132) | Men (n = 87) | P |
|---|---|---|---|
| **Demographics and exposures** |  |  |  |
| Age, years, mean (SD) | 47.3 (19.6); n=132 | 54.9 (16.5); n=87 | 0.002 |
| Body-mass index, kg/m<sup>2</sup>, mean (SD) | 25.9 (6.1); n=87 | 27.8 (6.4); n=66 | 0.034 |
| Admitted before 2012 | 55/132 (41.7%) | 16/87 (18.4%) | <0.001 |
| Oral contraception (women) | 52/132 (39.4%) | - |  |
| Pregnant or postpartum (women) | 5/132 (3.8%) | - |  |
| Other hormonal therapy | 16/132 (12.1%) | 4/87 (4.6%) | 0.091 |
| Any sex-specific risk factor (women) | 63/132 (47.7%) | - |  |
| Current smoking | 20/101 (19.8%) | 13/66 (19.7%) | 0.987 |
| Alcohol use | 42/83 (50.6%) | 39/59 (66.1%) | 0.066 |
| Previous venous thromboembolism | 18/132 (13.6%) | 21/87 (24.1%) | 0.047 |
| Active cancer | 18/132 (13.6%) | 7/87 (8.0%) | 0.203 |
| Hematologic disease | 15/132 (11.4%) | 8/87 (9.2%) | 0.609 |
| Chronic inflammatory/autoimmune disease | 15/132 (11.4%) | 7/87 (8.0%) | 0.424 |
| **Clinical presentation** |  |  |  |
| Symptom onset to admission, days, mean (SD) | 9.5 (16.5); n=117 | 9.1 (19.7); n=73 | 0.286 |
| Headache | 95/132 (72.0%) | 50/87 (57.5%) | 0.026 |
| with accompanying symptoms (among headache patients) | 42/95 (44.2%) | 12/50 (24.0%) | 0.017 |
| Focal neurological deficit | 87/132 (65.9%) | 53/87 (60.9%) | 0.452 |
| Acute symptomatic seizures (≤ 7 days) | 46/132 (34.8%) | 27/87 (31.0%) | 0.558 |
| mRS at admission, mean (SD) | 2.24 (1.59); n=131 | 1.84 (1.65); n=86 | 0.053 |
| NIHSS at admission, mean (SD) | 2.8 (4.6); n=118 | 2.2 (3.7); n=77 | 0.570 |
| GCS at admission, mean (SD) | 13.6 (3.0); n=129 | 14.0 (2.7); n=86 | 0.112 |
| **Imaging** |  |  |  |
| Number of thrombosed sinuses/veins, median [IQR] | 3 [1-3] | 2 [1-3] | 0.129 |
| ≥ 3 sites | 75/132 (56.8%) | 38/87 (43.7%) | 0.057 |
| Superior sagittal sinus | 63/132 (47.7%) | 43/87 (49.4%) | 0.806 |
| Transverse sinus | 91/132 (68.9%) | 62/87 (71.3%) | 0.714 |
| Sigmoid sinus | 82/132 (62.1%) | 50/87 (57.5%) | 0.491 |
| Internal jugular vein | 58/132 (43.9%) | 29/87 (33.3%) | 0.117 |
| Cortical veins | 19/132 (14.4%) | 5/87 (5.7%) | 0.045 |
| Deep venous system | 15/132 (11.4%) | 4/87 (4.6%) | 0.091 |
| Venous infarction | 46/132 (34.8%) | 23/87 (26.4%) | 0.190 |
| Congestive intracerebral hemorrhage | 52/132 (39.4%) | 25/87 (28.7%) | 0.106 |
| **Laboratory findings** |  |  |  |
| Systemic inflammation (composite) | 86/132 (65.2%) | 39/87 (44.8%) | 0.003 |
| elevated acute-phase markers only, no documented infection | 78/132 (59.1%) | 29/87 (33.3%) | <0.001 |
| documented infection | 8/132 (6.1%) | 7/87 (8.0%) | 0.569 |
| Elevated D-dimer (> 0.5 mg/L), among tested | 52/82 (63.4%) | 41/60 (68.3%) | 0.543 |
| Platelet count, ×10<sup>9</sup>/L, mean (SD) | 282 (101); n=130 | 217 (80); n=87 | <0.001 |
| **Treatment** |  |  |  |
| DOAC at discharge | 39/132 (29.5%) | 35/87 (40.2%) | 0.102 |
| VKA at discharge | 66/132 (50.0%) | 31/87 (35.6%) | 0.036 |
| Heparin only | 21/132 (15.9%) | 13/87 (14.9%) | 0.847 |
| No anticoagulation | 6/132 (4.5%) | 8/87 (9.2%) | 0.169 |
| Lifelong anticoagulation | 3/132 (2.3%) | 12/87 (13.8%) | 0.002 |
| Antiseizure medication at discharge | 45/132 (34.1%) | 26/87 (29.9%) | 0.515 |
| Hospital stay, days, mean (SD) | 12.3 (10.4); n=130 | 9.9 (7.4); n=86 | 0.022 |

Data are n/N (%) or mean (SD); the denominator is the number of patients with the item documented (Supplementary Table 1). Chi-squared or Fisher exact test for categorical variables; t-test for age and platelet count; Mann-Whitney U test for other continuous variables. Of the 40 comparisons in this table, six remained significant after Benjamini-Hochberg correction: platelet count, elevated acute-phase markers only, admission before 2012, lifelong anticoagulation, age and systemic inflammation (Supplementary Table 3). Sex-specific risk factor: oral contraception, pregnancy, puerperium or estrogen/progestogen therapy (corticosteroids and levothyroxine not counted). The two inflammation sub-rows do not sum to the composite in men: in 3 men the composite was positive with the source not specified in the registry (Supplementary Table 8). DOAC, direct oral anticoagulant; GCS, Glasgow Coma Scale; mRS, modified Rankin Scale; NIHSS, National Institutes of Health Stroke Scale; VKA, vitamin K antagonist.

### Presentation, imaging and laboratory findings

Headache was more frequent in women (72.0% versus 57.5%; P=0.026) and more often accompanied by nausea, photophobia or other symptoms (44.2% versus 24.0% of headache patients; P=0.017). Focal deficits, acute symptomatic seizures, admission NIHSS and GCS did not differ; admission mRS was numerically higher in women (2.24 versus 1.84; P=0.053) and hospital stay longer (12.3 versus 9.9 days; P=0.022) (Table 1).

Cortical vein thrombosis was more frequent in women (14.4% versus 5.7%; P=0.045). Involvement of the deep venous system (11.4% versus 4.6%; P=0.091) and of three or more sites (56.8% versus 43.7%; P=0.057) was numerically more frequent in women. The median number of thrombosed sinuses and veins was 3 [IQR 1-3] in women and 2 [1-3] in men (P=0.13; Table 1, Supplementary Table 7). Venous infarction (34.8% versus 26.4%) and congestive intracerebral hemorrhage (39.4% versus 28.7%) did not differ significantly.

The inflammation composite was positive in 65.2% of women and 44.8% of men (P=0.003). The difference was confined to the markers-only phenotype (59.1% versus 33.3%; P<0.001); documented infections (6.1% versus 8.0%) and chronic inflammatory disease (11.4% versus 8.0%) were equally frequent. Elevated CRP (51.5% versus 34.5%; P=0.013) and leukocytosis (35.6% versus 21.8%; P=0.030) were each more often documented in women (Supplementary

Table 8). The composite was more often positive in women both with and without a sex-specific risk factor (68.3% and 62.3% versus 44.8% in men; Supplementary Table 6). The markers-only phenotype was associated with parenchymal lesions and acute symptomatic seizures and unrelated to oral contraception, D-dimer or 6-month outcome (Supplementary Table 8). Platelet counts were higher in women (282 ± 101 versus 217 ± 80 × 10⁹/L; P<0.001); an elevated D-dimer was equally frequent among those tested (63.4% versus 68.3%).

Of the 40 unadjusted comparisons in Table 1, six remained significant after Benjamini-Hochberg correction: platelet count, elevated acute-phase markers without documented infection, admission before 2012, lifelong anticoagulation, age and the inflammation composite (Supplementary Table 3).

### Sex differences after adjustment for age and body-mass index

After adjustment for age the association of female sex with headache was attenuated (odds ratio [OR], 1.42 [95% CI, 0.74-2.74]). After adjustment for age and BMI it was 1.17 (0.54-2.54) in the 153 patients with BMI recorded and 1.53 (0.78-3.00) in all 219 patients with BMI imputed (Table 2). The association with cortical vein thrombosis was attenuated in the same way (age-adjusted 2.38, 0.83-6.77; age- and BMI-adjusted 2.05, 0.59-7.06; BMI imputed 2.57, 0.88-7.47), as was the longer hospital stay (+1.7 days, −1.2 to +4.6). The associations with the inflammation composite (age- and BMI-adjusted OR, 2.29 [95% CI, 1.16-4.50]; BMI imputed 2.31, 1.30-4.11), with the markers-only phenotype (2.91, 1.44-5.87; imputed 2.90, 1.61-5.22) and with the platelet count (+56 × 10⁹/L, +26 to +86; imputed +63, +37 to +89) persisted. Adding calendar period changed none of these estimates materially (composite 2.22, 1.12-4.39; platelets +55 × 10⁹/L; Supplementary Table 9). Adding the number of thrombosed sites did not change the estimates for headache (1.41, 0.73-2.72), hemorrhage (1.61, 0.89-2.91) or venous infarction (1.63, 0.88- 3.00) (Supplementary Table 10). Women more often received a vitamin K antagonist at discharge (50.0% versus 35.6%; P=0.036), an association that disappeared after adjustment for calendar period (OR, 0.97 [95% CI, 0.49-1.94]; Supplementary Table 9).

**Table 2.**
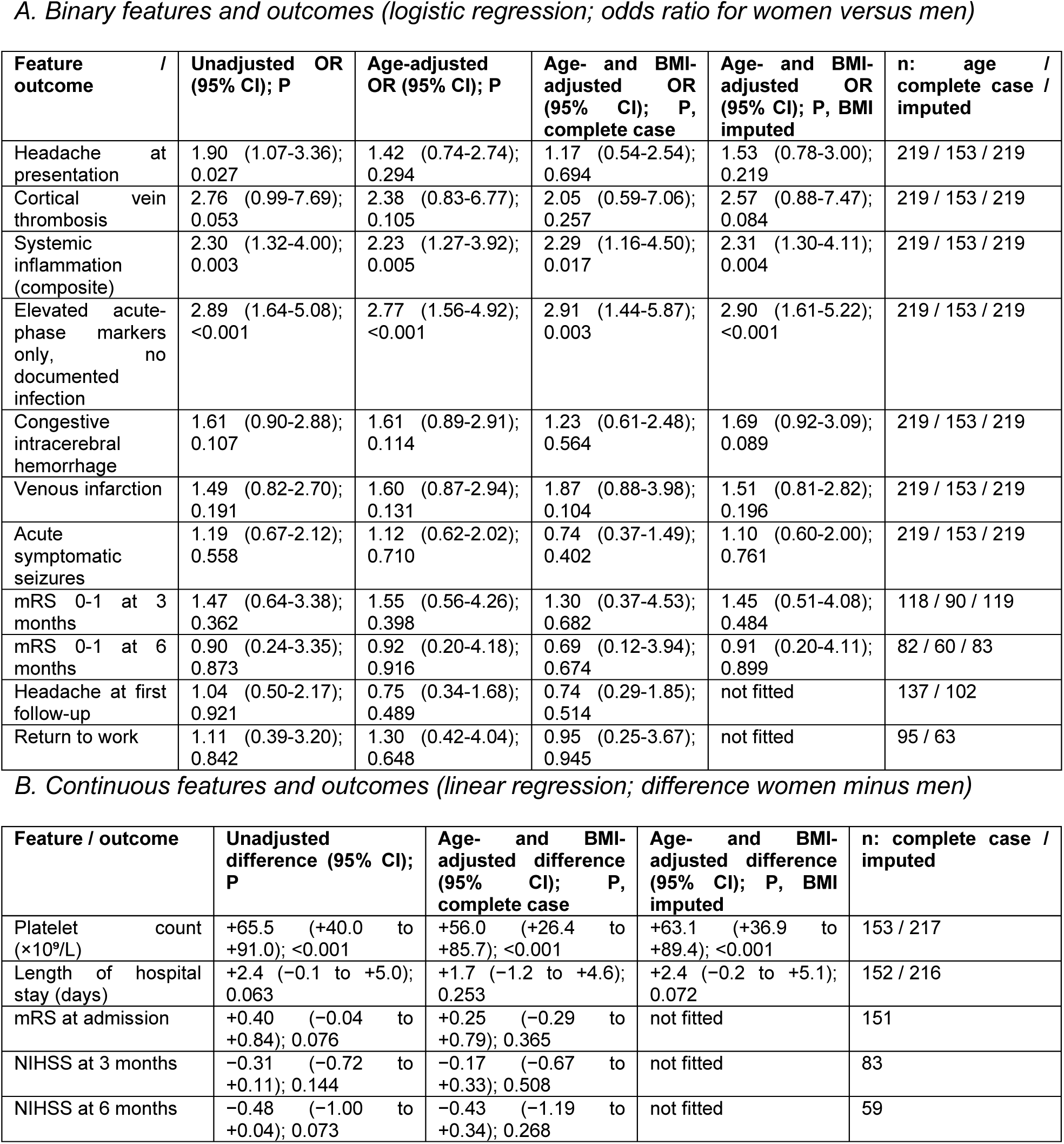
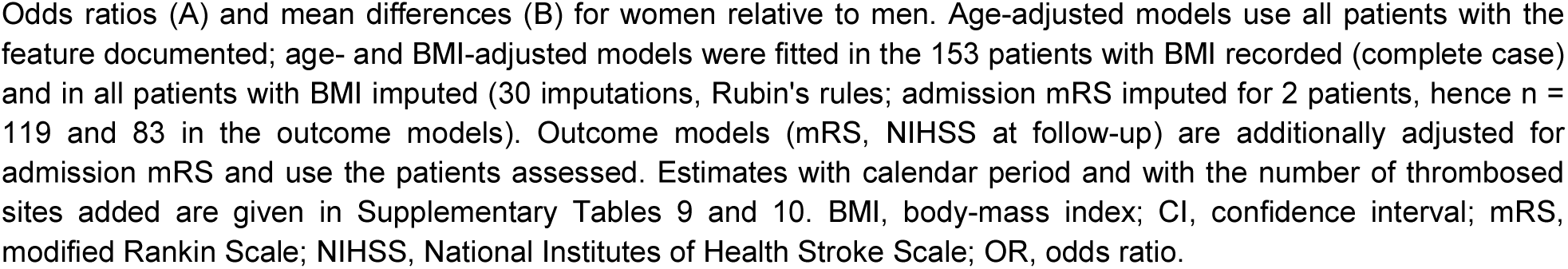
Sex differences after adjustment for age and body-mass index.

| *A. Binary features and outcomes (logistic regression; odds ratio for women versus men)* |  |  |  |  |  |
|---|---|---|---|---|---|
| **Feature / outcome** | **Unadjusted OR (95% CI); P** | **Age-adjusted OR (95% CI); P** | **Age- and BMI-adjusted OR (95% CI); P, complete case** | **Age- and BMI-adjusted OR (95% CI); P, BMI imputed** | **n: age / complete case / imputed** |
| Headache at presentation | 1.90 (1.07-3.36); 0.027 | 1.42 (0.74-2.74); 0.294 | 1.17 (0.54-2.54); 0.694 | 1.53 (0.78-3.00); 0.219 | 219 / 153 / 219 |
| Cortical vein thrombosis | 2.76 (0.99-7.69); 0.053 | 2.38 (0.83-6.77); 0.105 | 2.05 (0.59-7.06); 0.257 | 2.57 (0.88-7.47); 0.084 | 219 / 153 / 219 |
| Systemic inflammation (composite) | 2.30 (1.32-4.00); 0.003 | 2.23 (1.27-3.92); 0.005 | 2.29 (1.16-4.50); 0.017 | 2.31 (1.30-4.11); 0.004 | 219 / 153 / 219 |
| Elevated acute-phase markers only, no documented infection | 2.89 (1.64-5.08); <0.001 | 2.77 (1.56-4.92); <0.001 | 2.91 (1.44-5.87); 0.003 | 2.90 (1.61-5.22); <0.001 | 219 / 153 / 219 |
| Congestive intracerebral hemorrhage | 1.61 (0.90-2.88); 0.107 | 1.61 (0.89-2.91); 0.114 | 1.23 (0.61-2.48); 0.564 | 1.69 (0.92-3.09); 0.089 | 219 / 153 / 219 |
| Venous infarction | 1.49 (0.82-2.70); 0.191 | 1.60 (0.87-2.94); 0.131 | 1.87 (0.88-3.98); 0.104 | 1.51 (0.81-2.82); 0.196 | 219 / 153 / 219 |
| Acute symptomatic seizures | 1.19 (0.67-2.12); 0.558 | 1.12 (0.62-2.02); 0.710 | 0.74 (0.37-1.49); 0.402 | 1.10 (0.60-2.00); 0.761 | 219 / 153 / 219 |
| mRS 0-1 at 3 months | 1.47 (0.64-3.38); 0.362 | 1.55 (0.56-4.26); 0.398 | 1.30 (0.37-4.53); 0.682 | 1.45 (0.51-4.08); 0.484 | 118 / 90 / 119 |
| mRS 0-1 at 6 months | 0.90 (0.24-3.35); 0.873 | 0.92 (0.20-4.18); 0.916 | 0.69 (0.12-3.94); 0.674 | 0.91 (0.20-4.11); 0.899 | 82 / 60 / 83 |
| Headache at first follow-up | 1.04 (0.50-2.17); 0.921 | 0.75 (0.34-1.68); 0.489 | 0.74 (0.29-1.85); 0.514 | not fitted | 137 / 102 |
| Return to work | 1.11 (0.39-3.20); 0.842 | 1.30 (0.42-4.04); 0.648 | 0.95 (0.25-3.67); 0.945 | not fitted | 95 / 63 |
| *B. Continuous features and outcomes (linear regression; difference women minus men)* |  |  |  |  |  |
| **Feature / outcome** | **Unadjusted difference (95% CI); P** | **Age- and BMI-adjusted difference (95% CI); P, complete case** | **Age- and BMI-adjusted difference (95% CI); P, BMI imputed** | **n: complete case / imputed** |  |
| Platelet count (×10<sup>9</sup>/L) | +65.5 (+40.0 to +91.0); <0.001 | +56.0 (+26.4 to +85.7); <0.001 | +63.1 (+36.9 to +89.4); <0.001 | 153 / 217 |  |
| Length of hospital stay (days) | +2.4 (−0.1 to +5.0); 0.063 | +1.7 (−1.2 to +4.6); 0.253 | +2.4 (−0.2 to +5.1); 0.072 | 152 / 216 |  |
| mRS at admission | +0.40 (−0.04 to +0.84); 0.076 | +0.25 (−0.29 to +0.79); 0.365 | not fitted | 151 |  |
| NIHSS at 3 months | −0.31 (−0.72 to +0.11); 0.144 | −0.17 (−0.67 to +0.33); 0.508 | not fitted | 83 |  |
| NIHSS at 6 months | −0.48 (−1.00 to +0.04); 0.073 | −0.43 (−1.19 to +0.34); 0.268 | not fitted | 59 |  |

Odds ratios (A) and mean differences (B) for women relative to men. Age-adjusted models use all patients with the feature documented; age- and BMI-adjusted models were fitted in the 153 patients with BMI recorded (complete case) and in all patients with BMI imputed (30 imputations, Rubin's rules; admission mRS imputed for 2 patients, hence n = 119 and 83 in the outcome models). Outcome models (mRS, NIHSS at follow-up) are additionally adjusted for admission mRS and use the patients assessed. Estimates with calendar period and with the number of thrombosed sites added are given in Supplementary Tables 9 and 10. BMI, body-mass index; CI, confidence interval; mRS, modified Rankin Scale; NIHSS, National Institutes of Health Stroke Scale; OR, odds ratio.

### Follow-up and functional outcome by sex

A 3-month mRS was documented in 119/219 patients (54.3%), a 6-month mRS in 83 (37.9%) and a 12-month visit in 47 (21.5%); median clinical follow-up was 0.51 years (IQR 0.08-1.80; Supplementary Table 11). Assessment at 3 and 6 months did not differ by sex (51.5% versus 58.6% and 38.6% versus 36.8%); a 12-month visit was more often documented in women (26.5% versus 13.8%; P=0.025; adjusted OR, 2.03 [95% CI, 0.92-4.46]). Assessment was less complete before 2012 and in older patients and more complete in patients with headache, seizures or a direct oral anticoagulant at discharge, and did not depend on admission severity (Supplementary Tables 12-13, Supplementary Figure 2).

At 3 months, 77.9% of assessed women and 70.6% of assessed men had mRS 0-1 (P=0.36; adjusted OR, 1.30 [95% CI, 0.37-4.53]), and at 6 months 86.3% and 87.5% of assessed survivors (adjusted OR, 0.69 [95% CI, 0.12-3.94]; Table 3, Figure 1). Fifteen patients (9 women, 6 men) were known to have died before the 6-month visit. For the whole cohort, the estimated proportion with mRS 0-1 at 6 months was 74.1% (95% CI, 67.2-80.9) by multiple imputation and 72.1% by inverse-probability weighting, with extreme-case bounds of 32.9% and 88.1%. A 20-point shortfall in unassessed survivors would lower the estimate to 63.9% (Supplementary Table 14). NIHSS at 6 months was lower in women (0.14 ± 0.72 versus 0.61 ± 1.63; Mann-Whitney P=0.025), a difference in the proportion with a score of exactly zero (94.1% versus 77.4%). The mean difference was −0.48 points (95% CI, −1.00 to +0.04) unadjusted, −0.43 (−1.19 to +0.34) after adjustment and −0.27 (−0.78 to +0.24) after multiple imputation of the missing 6-month scores (Supplementary Table 14). Within women, functional outcome did not differ by sex-specific risk factor (mRS 0-1 at 6 months 80.6% with versus 95.0% without; Supplementary Table 6). Death from any cause during observation occurred in 11.4% of women and 17.2% of men (P=0.22); the four deaths attributed to CVT were all in women. Recurrent CVT occurred in one woman and one man among 141 patients with documented status.

**Figure 1.**
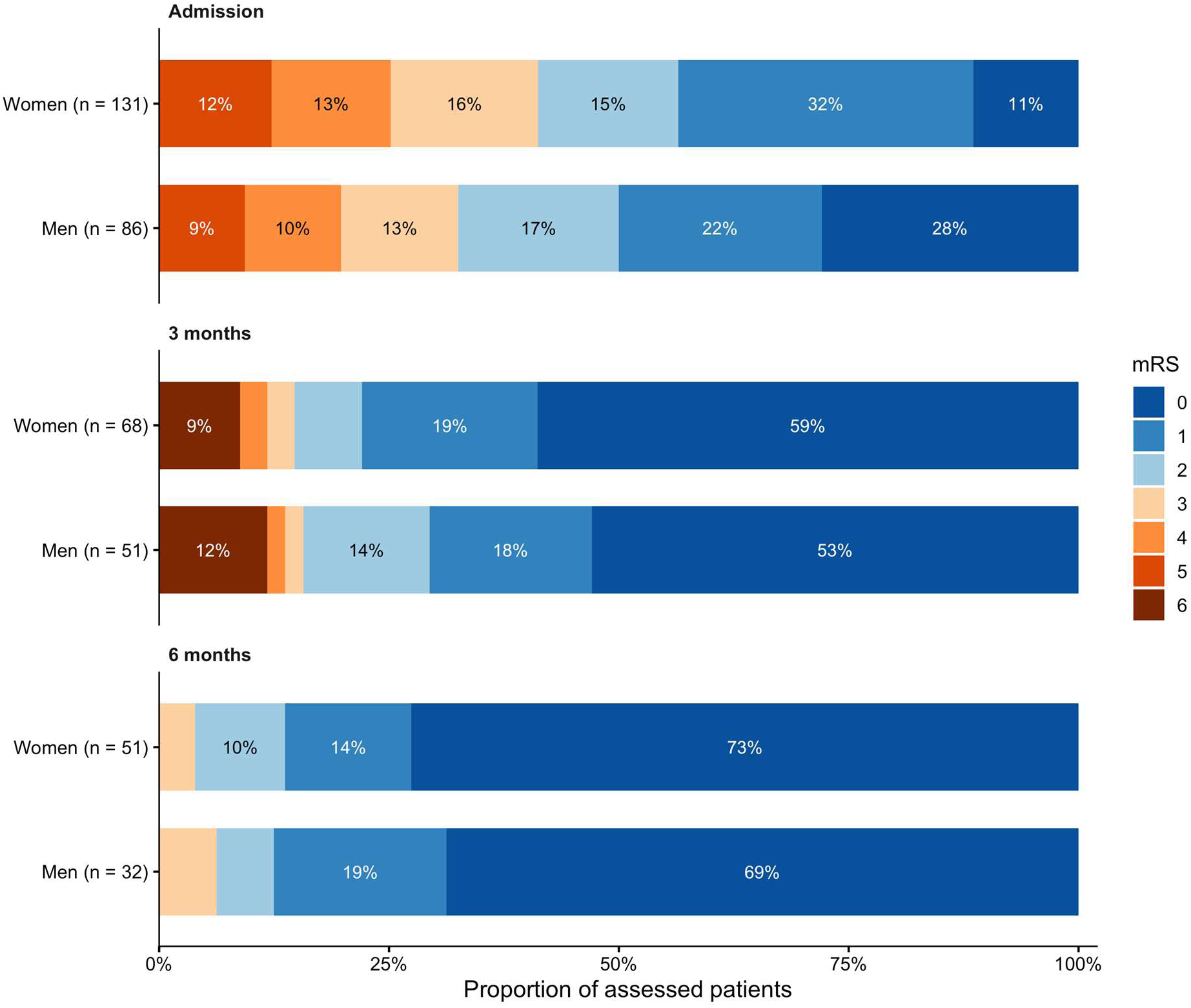
Distribution of the modified Rankin Scale by sex at admission, 3 months and 6 months. Stacked bars give the percentage of patients in each mRS category among those assessed (n in the axis labels); at admission and 3 months the distributions include patients recorded as mRS 6 (at 3 months, 6 women and 6 men), and the 6-month mRS was recorded for survivors only. Segment labels give percentages (omitted below 8%). mRS, modified Rankin Scale.

**Table 3.** Follow-up, functional outcome and residual symptoms by sex.

|  | **All patients (n = 219)** | **Women (n = 132)** | **Men (n = 87)** |
|---|---|---|---|
| **3-month follow-up** |  |  |  |
| mRS assessed, n/N (%) | 119/219 (54.3%) | 68/132 (51.5%) | 51/87 (58.6%) |
| mRS 0-1 | 89/119 (74.8%) | 53/68 (77.9%) | 36/51 (70.6%) |
| mRS 6 (death) | 12/119 (10.1%) | 6/68 (8.8%) | 6/51 (11.8%) |
| NIHSS, mean (SD) | 0.31 (1.06); n=105 | 0.18 (0.57); n=60 | 0.49 (1.47); n=45 |
| **6-month follow-up** |  |  |  |
| mRS assessed (survivors), n/N (%) | 83/219 (37.9%) | 51/132 (38.6%) | 32/87 (36.8%) |
| mRS 0-1 | 72/83 (86.7%) | 44/51 (86.3%) | 28/32 (87.5%) |
| Died before the 6-month assessment | 15/219 (6.8%) | 9/132 (6.8%) | 6/87 (6.9%) |
| NIHSS, mean (SD) | 0.32 (1.16); n=82 | 0.14 (0.72); n=51 | 0.61 (1.63); n=31 |
| 12-month visit documented, n/N (%) | 47/219 (21.5%) | 35/132 (26.5%) | 12/87 (13.8%) |
| **First follow-up with the item documented** |  |  |  |
| Persistent headache | 43/137 (31.4%) | 26/82 (31.7%) | 17/55 (30.9%) |
| Seizure-free | 134/143 (93.7%) | 83/87 (95.4%) | 51/56 (91.1%) |
| Antiseizure medication (status documented) | 51/57 (89.5%) | 33/35 (94.3%) | 18/22 (81.8%) |
| Returned to work (employed patients) | 77/95 (81.1%) | 49/60 (81.7%) | 28/35 (80.0%) |
| **Among patients with mRS 0-1 at 6 months** |  |  |  |
| Persistent headache | 26/66 (39.4%) | 16/41 (39.0%) | 10/25 (40.0%) |
| Not returned to work | 7/51 (13.7%) | 5/34 (14.7%) | 2/17 (11.8%) |
| On antiseizure medication (seizure patients) | 23/24 (95.8%) | 15/15 (100.0%) | 8/9 (88.9%) |
| Headache or not back at work | 30/69 (43.5%) | 18/43 (41.9%) | 12/26 (46.2%) |
| **Events during observation** |  |  |  |
| Recurrent CVT (status documented) | 2/141 (1.4%) | 1/84 (1.2%) | 1/57 (1.8%) |
| Death attributed to CVT | 4/219 (1.8%) | 4/132 (3.0%) | 0/87 (0.0%) |
| Death, any cause | 30/219 (13.7%) | 15/132 (11.4%) | 15/87 (17.2%) |
| **Recanalization on follow-up imaging** |  |  |  |
| Recanalization documented (complete or partial), whole cohort | 77/219 (35.2%) | 49/132 (37.1%) | 28/87 (32.2%) |
| complete recanalization at the 6-month scan (among imaged) | 51/95 (53.7%) | 33/59 (55.9%) | 18/36 (50.0%) |
| complete recanalization at the 12-month visit (among imaged) | 59/86 (68.6%) | 39/53 (73.6%) | 20/33 (60.6%) |

Data are n/N (%) or mean (SD); the denominator of every proportion is the number of patients with the item documented. The 6-month mRS was recorded for survivors; the row below gives the patients known to have died before that visit. Headache, seizure status, antiseizure medication and return to work are undated registry items and refer to the first follow-up at which they were documented. Recanalization at the 6-month scan and the 12-month visit is given among the patients imaged at that visit; Supplementary Table 16 gives the cumulative incidence of first documented complete or partial recanalization among imaged patients with a datable observation, censored at the last documented imaging assessment. P values for the sex comparisons are given in Supplementary Table 15; none was < 0.05 except the 12-month visit (P=0.025) and the 6-month NIHSS (P=0.025). CVT, cerebral venous thrombosis; mRS, modified Rankin Scale; NIHSS, National Institutes of Health Stroke Scale.

### Residual symptoms and return to work by sex

At the first follow-up at which the item was documented, persistent headache was reported by 31.7% of women and 30.9% of men, and 81.7% and 80.0% of previously employed patients had returned to work. Seizure freedom was documented in 95.4% and 91.1% (Table 3). Among the 72 patients with mRS 0-1 at 6 months, 39.0% of women and 40.0% of men with the item documented reported persistent headache, 14.7% and 11.8% had not returned to work, and 15 of 15 and 8 of 9 seizure patients remained on ASM. Persistent headache or failure to return to work was present in 41.9% of women and 46.2% of men (P=0.73; Supplementary Table 15). Neither persistent headache nor return to work differed by sex after adjustment for age and BMI (Table 2).

### Recanalization by sex

Among the 169 imaged patients with a datable observation, the cumulative incidence of documented recanalization (complete or partial) was 58.4% in women and 49.8% in men at 6 months, and 73.8% (95% CI, 60.9-85.2) and 56.9% (42.1-72.7) at 12 months. The difference was compatible with chance (log-rank P=0.37; HR for women adjusted for age and BMI, 1.06 [95% CI, 0.59-1.87] in the 122 patients with BMI recorded and 0.97 [95% CI, 0.58-1.61] with BMI imputed; Supplementary Table 16). Complete recanalization at the 6-month scan was documented in 55.9% of imaged women and 50.0% of imaged men (Table 3).

## Discussion

This single-center cohort of 219 consecutive adults with CVT treated between 2001 and 2021 compares women and men at equal age and BMI. It separates sex differences accounted for by age and BMI from those that persist after adjustment. The female excess of headache, which earlier cohorts reported without adjustment, was attenuated once age was accounted for and became compatible with chance after adjustment for age and BMI; the excess of cortical vein thrombosis was imprecise even before adjustment (Table 2). The higher platelet count and the more frequent elevation of acute-phase markers without an infectious focus in women persisted after adjustment for age, BMI and calendar period. Neither kind of difference was accompanied by a difference in functional outcome, residual symptoms, return to work or recanalization among the patients assessed.

Our patients were older than those in ISCVT, VENOST and CEVETIS (mean 50 years versus reported means or medians of 34-42 years),^4,5,7^ women accounted for a smaller proportion of the cohort (60% versus 63-75%),^4,5,7,13^ and fewer women had a sex-specific risk factor (48% versus 65% in ISCVT).^4^ Focal deficits (64%) and acute symptomatic seizures (33%) were within the range of earlier cohorts.^2,4,7^

Women with a sex-specific risk factor (oral contraception, pregnancy, puerperium or estrogen or progestogen therapy) were younger and more often presented with headache (mean age 34 years; headache in 90%), whereas women without these factors resembled men (mean age 59 versus 55 years; headache in 55% versus 58%; Supplementary Table 6). In ISCVT, VENOST and CEVETIS, women were younger than men and presentation was compared without adjustment for age^4,5,7^; the ISCVT authors considered the higher rate of headache in women with a sex-specific risk factor a possible effect of age,^4^ and CEVETIS did not compare presenting symptoms.^5^ In our cohort the OR for headache in women fell from 1.9 to 1.4 with adjustment for age, and to 1.2 in the complete-case and 1.5 in the imputed analysis with BMI added; the confidence intervals include no difference and a moderate female excess alike. Cortical vein thrombosis, more frequent in women in CEVETIS (6.0% versus 2.8%) as it was here (14.4% versus 5.7%),^5^ behaved in the same way. Age and sex-specific risk factors therefore contribute to the differences in presentation between women and men described across cohorts. The Vellore registry makes the same point from the other direction: as postpartum CVT declined, the sex ratio reversed and the mean age rose.^11^

Two differences persisted after adjustment for age, BMI and calendar period and may reflect sex-specific patterns: the inflammation composite and the platelet count. The composite was more often positive in women both with and without a sex-specific risk factor (68% and 62% versus 45% in men). In 86% of patients with a positive composite, elevated CRP or leukocyte counts were documented without an infectious focus, and this markers-only phenotype was associated with parenchymal lesions and acute symptomatic seizures but not with oral contraception. One possible explanation is a non-specific acute-phase response to venous congestion and parenchymal injury that is more marked in women. A higher erythrocyte sedimentation rate in women was reported in one series,^12^ whereas VENOST found no sex difference in inflammatory markers.^7^ In our registry, markers were recorded as elevated or not, with no category for a test not performed, so more frequent testing in women would produce the same pattern.

Platelet counts in women were about 60 × 10⁹/L higher after adjustment, three times the physiological sex difference of about 20 × 10⁹/L, which narrows with age^17^; reactive changes are an alternative explanation. Anemia, which is associated with CVT and with reactive thrombocytosis,^18^ is another possible contributor, but hemoglobin values were not available in the registry. Neither finding predicted outcome in this cohort, and we present both as hypotheses for prospective cohorts with complete laboratory data.

Pregnancy- or puerperium-associated CVT was rare (3.8% of women, versus 10.6% in CEVETIS and 20-50% in Vellore).^5,11^ The most likely explanation is case identification through neurology department records: pregnancy-associated CVT at our center is partly managed by obstetrics and neurosurgery with neurological consultation, and such patients were not captured.

This is a limitation for the sex-specific aims of the study, because the youngest women with the most characteristic hormonal risk profile are under-represented, and it may contribute to the older age of our women (mean 47 years) compared with the registries. The direction of the resulting bias is toward fewer sex differences in presentation and toward a smaller female advantage in outcome, since young women with a sex-specific risk factor had the best outcome in ISCVT.^4^

Functional outcome among assessed patients did not differ by sex (86% and 88% independent at 6 months); neither did return to work, residual headache or seizure freedom. The better outcome of women with a sex-specific risk factor reported from ISCVT^4^ was not apparent here (81% versus 95% in women without and 88% in men; Supplementary Table 6). The lower 6- month NIHSS in women reflects more scores of zero and is compatible with chance after adjustment. These comparisons rest on 83 patients with a 6-month mRS; the confidence intervals of the adjusted ORs include at least a four-fold difference in either direction.

Within the 72 patients independent at 6 months, four in ten with documented headache status reported persistent headache, one in seven with occupational data had not returned to work, and nearly all seizure patients with medication data remained on ASM; 44% had persistent headache or had not returned to work, equally in women and men (42% and 46%). These items were recorded during follow-up and not necessarily at the 6-month visit. Cohorts with detailed follow-up found residual symptoms in 42% despite mRS 0-1 in 84%,^19^ and concentration impairment in 75% and headache in 43% of functionally independent patients after five years.^20^ With 79% of ISCVT patients^2^ and 87% of our assessed survivors at mRS 0-1, outcomes cluster at the favorable end of the scale, and mRS-based comparisons reveal little about residual morbidity in either sex. In a Swedish cohort, female sex was associated with not returning to work^6^; this was not reproduced, although our occupational item was undated and recorded in 95 patients only. Follow-up after CVT should record headache, cognition, fatigue, mood and return to work with patient-reported instruments and review the ongoing indication for ASM.

Recanalization did not differ detectably by sex (73.8% versus 56.9% at 12 months, log-rank P=0.37), consistent with the CEVETIS report that sex was unrelated to the degree of recanalization.^5^ Our estimate is conditional on imaging having been performed and on a registry code that does not separate absent from unknown recanalization, and is presented descriptively.

### Strengths and limitations

Strengths are the consecutive inclusion of venographically confirmed cases over 21 years, the re-estimation of every sex difference at equal age and BMI, and a script that reproduces every reported number. The limitations are those of retrospective data from a single center. The mRS was documented in 54% of patients at 3 months and 38% at 6 months. Assessment was less complete before 2012 and in older patients but did not depend on sex or on recorded admission severity, and the whole-cohort estimates from imputation and weighting (74% and 72%) lie within a few points of the proportion among patients with a known 6-month status, deaths included (74%). Sex comparisons of outcome within assessed patients are therefore less exposed to selection than whole-cohort estimates, but they are not protected from it, and their precision is low.

Presenting symptoms and inflammatory markers were coded as present or absent without a category for an item not assessed, so differences in documentation between the sexes cannot be excluded, although the documented laboratory items did not differ by sex. BMI was recorded in 153 patients; the imputed and complete-case estimates agreed, which argues against a large selection effect of the BMI subset. Because age and BMI may lie on the causal pathway between sex and presentation, the adjusted models describe sex differences at equal age and BMI and do not estimate a total effect of sex. No patient had a pre-existing headache disorder among the documented comorbidities, which were abstracted from the admission letters; under-documentation cannot be excluded. The occupational and symptom items were undated and measured without instruments; misclassification that did not depend on sex would bias these comparisons toward no difference. Patients with CVT treated in other departments were not captured, which may have under-ascertained pregnancy-associated CVT. Recruitment over 21 years introduces heterogeneity in imaging, treatment and follow-up practice, which we addressed by adding calendar period to the models and by presenting sex differences by period (Supplementary Table 9); the period adjustment did not alter the conclusions. Forty unadjusted comparisons were made in Table 1; six survived correction for multiplicity, among them the two differences that also persisted after adjustment for age and BMI, the platelet count and the elevation of acute-phase markers without documented infection.

## Conclusions

At equal age and BMI, women and men with CVT differed less in presentation than the unadjusted comparisons of earlier cohorts suggest. The female excess of headache and cortical vein thrombosis was attenuated, whereas a higher platelet count and a non-specific elevation of acute-phase markers in women persisted. Neither difference was accompanied by a difference in functional outcome, residual symptoms or return to work among the patients assessed. These conclusions rest on a cohort with incomplete follow-up; the persisting laboratory differences should be read as hypotheses for prospective cohorts with complete laboratory data.

## Data Availability

Data sharing statement. The individual participant data underlying this article are part of an ongoing registry study and cannot be shared, in accordance with the terms of the ethics approval (KEK 2021-01558) and Swiss data-protection law. The analysis code is provided as supplementary material. Requests concerning the data or methods may be directed to the corresponding author.

## Declarations

## Data sharing statement

The individual participant data underlying this article are part of an ongoing registry study and cannot be shared, in accordance with the terms of the ethics approval (KEK 2021-01558) and Swiss data-protection law. The analysis code is provided as supplementary material. Requests concerning the data or methods may be directed to the corresponding author.

## Acknowledgements

We thank all patients included in the study, all of whom provided general consent for use of their data for research purposes. Generative artificial-intelligence tools (Claude, Anthropic; GPT, OpenAI) were used under the direction and supervision of the authors for language and analysis code editing, and for automated consistency checks between the manuscript and the analysis output; the study design, data abstraction and verification, the choice and interpretation of the analyses and the final text are the work of the authors, who take full responsibility for the content.

## Ethical approval

Ethics Committee of the Canton of Zurich (KEK 2021-01558). All included patients had provided general consent.

## Author contributions

Conceptualization: TIG, MB, MK, SW, AS. Methodology: TIG, MB, MK, SW, AS. Investigation: TIG, MR. Data curation: TIG, MR. Formal analysis: TIG, AS, LH. Validation: TIG, AS. Writing - original draft: TIG, AS. Writing - review & editing: MR, LH, MB, MK, PT, ZK, AL, MW, SW. All authors contributed to the interpretation of the data, revised the manuscript critically for important intellectual content and approved the final version. TIG and AS had full access to all data in the study, directly accessed and verified the underlying data, and take responsibility for the integrity of the data and the accuracy of the analysis.

## References

1. Saposnik G, Bushnell C, Coutinho JM, et al. Diagnosis and management of cerebral venous thrombosis: a scientific statement from the American Heart Association. Stroke. 2024;55:e77–e90. doi:10.1161/STR.0000000000000456

2. Ferro JM, Canhão P, Stam J, Bousser MG, Barinagarrementeria F. Prognosis of cerebral vein and dural sinus thrombosis: results of the International Study on Cerebral Vein and Dural Sinus Thrombosis (ISCVT). Stroke. 2004;35:664–670. doi:10.1161/01.STR.0000117571.76197.26

3. Silvis SM, Lindgren E, Hiltunen S, et al. Postpartum period is a risk factor for cerebral venous thrombosis. Stroke. 2019;50:501–503. doi:10.1161/STROKEAHA.118.023017

4. Coutinho JM, Ferro JM, Canhão P, et al. Cerebral venous and sinus thrombosis in women. Stroke. 2009;40:2356–2361. doi:10.1161/STROKEAHA.108.543884

5. Porceddu E, Rezoagli E, Poli D, et al. Sex-related characteristics of cerebral vein thrombosis: a secondary analysis of a multicenter international cohort study. Thromb Res. 2020;196:371–374. doi:10.1016/j.thromres.2020.09.023

6. Lindgren E, Jood K, Tatlisumak T. Vocational outcome in cerebral venous thrombosis: long-term follow-up study. Acta Neurol Scand. 2018;137:299–307. doi:10.1111/ane.12875

7. Duman T, Uluduz D, Midi I, et al. A multicenter study of 1,144 patients with cerebral venous thrombosis: the VENOST study. J Stroke Cerebrovasc Dis. 2017;26:1848–1857. doi:10.1016/j.jstrokecerebrovasdis.2017.04.020

8. Ranjan R, Ken-Dror G, Martinelli I, et al. Age of onset of cerebral venous thrombosis: the BEAST study. Eur Stroke J. 2023;8:344–350. doi:10.1177/23969873221148267

9. Zuurbier SM, Hiltunen S, Lindgren E, et al. Cerebral venous thrombosis in older patients. Stroke. 2018;49:197–200. doi:10.1161/STROKEAHA.117.019483

10. Zuurbier SM, Arnold M, Middeldorp S, et al. Risk of cerebral venous thrombosis in obese women. JAMA Neurol. 2016;73:579–584. doi:10.1001/jamaneurol.2016.0001

11. Miraclin TA, Prasad JD, Ninan GA, et al. Cerebral venous sinus thrombosis: changing trends in the incidence, age and gender (findings from the CMC Vellore CVT registry). Stroke Vasc Neurol. 2024;9:252–257. doi:10.1136/svn-2023-002351

12. Zarimeidani F, Rahmati R, Mokary Y, et al. Sex differences in clinical profile and risk factors of cerebral venous sinus thrombosis at a high altitude area. Medicine (Baltimore). 2024;103:e38711. doi:10.1097/MD.0000000000038711

13. Bajko Z, Motataianu A, Stoian A, et al. Gender differences in risk factor profile and clinical characteristics in 89 consecutive cases of cerebral venous thrombosis. J Clin Med. 2021;10:1382. doi:10.3390/jcm10071382

14. von Elm E, Altman DG, Egger M, Pocock SJ, Gøtzsche PC, Vandenbroucke JP. The Strengthening the Reporting of Observational Studies in Epidemiology (STROBE) statement: guidelines for reporting observational studies. Lancet. 2007;370:1453–1457. doi:10.1016/S0140-6736(07)61602-X

15. Heidari S, Babor TF, De Castro P, Tort S, Curno M. Sex and Gender Equity in Research: rationale for the SAGER guidelines and recommended use. Res Integr Peer Rev. 2016;1:2. doi:10.1186/s41073-016-0007-6

16. Beghi E, Carpio A, Forsgren L, et al. Recommendation for a definition of acute symptomatic seizure. Epilepsia. 2010;51:671–675. doi:10.1111/j.1528-1167.2009.02285.x

17. Balduini CL, Noris P. Platelet count and aging. Haematologica. 2014;99:953–955. doi:10.3324/haematol.2014.106260

18. Coutinho JM, Zuurbier SM, Gaartman AE, et al. Association between anemia and cerebral venous thrombosis: case-control study. Stroke. 2015;46:2735–2740. doi:10.1161/STROKEAHA.115.009843

19. Hiltunen S, Putaala J, Haapaniemi E, Tatlisumak T. Long-term outcome after cerebral venous thrombosis: analysis of functional and vocational outcome, residual symptoms, and adverse events in 161 patients. J Neurol. 2016;263:477–484. doi:10.1007/s00415-015-7996-9

20. Koopman K, Uyttenboogaart M, Vroomen PC, van der Meer J, De Keyser J, Luijckx GJ. Long-term sequelae after cerebral venous thrombosis in functionally independent patients. J Stroke Cerebrovasc Dis. 2009;18:198–202. doi:10.1016/j.jstrokecerebrovasdis.2008.10.004

